# Adiposity and lean muscle indices that associate with cardiometabolic risk factors and inflammatory mediators in a South Asian population

**DOI:** 10.1101/2025.04.13.25325724

**Authors:** Dumni Nadani Gunasinghe, Jeewantha Jayamali, Padukkage Harshani Chathurangika, Thashmi Nimasha, Lahiru Perera, Uvini Amarasekara, Rivindu H. Wickramanayake, Aayuka Panditharatne, Gathsaurie Neelika Malavige, Chandima Jeewandara

## Abstract

**Background:** Cardiovascular diseases (CVD) are the leading cause of death in South Asia with visceral adiposity and sarcopenic obesity emerging as critical risk factors.

**Methods:** We conducted a cross-sectional study in 139 Sri Lankan adults (mean age 39.3, SD ± 10.7, 52 females and 87 males) to assess visceral adipose tissue (VAT) area, fat mass index (FMI), fat mass ratio (FMR) and lean muscle mass, expressed as the appendicular lean mass index (ALMI) using dual energy Xray absorptiometry (DEXA). Fasting blood sugar (FBS, lipid profiles, apolipoproteins (A1 and B) and inflammatory markers (IL-6 and IL-1b) were analysed.

**Results:** 23 (44%) females and 67 (77%) of males had a VAT area of ≥ 100cm^2^, 34 (65%) females and 79 (91%) males had an FMI of above the cut-off value and 30 (58%) females, and 33 (38%) males had an FMR above the cut-off value. Females with high VAT area (≥100cm^2^) had significantly higher levels of FBS (p=0.008) and lower HDL and Apo A1 (p<0.05) compared to females with a VAT area of <100cm^2^. Further, females with an FMI of ≥9 had significantly higher FBS levels (p=0.01), and those with a FMR of ≥ of 1.2 were had significantly higher FBS and ApoB levels with significantly lower ApoA1 levels. There were no differences in metabolic risk factors or inflammatory mediators in males with an FMI above the cut-off value (≥6) or with a high FMR. 34 (65%) females had a low ALMI below (<5.5), while 47 (54%) males had a low ALMI (<7). ApoB levels, ApoB/ApoA1 ratio and serum cholesterol were significantly higher in males with a ALMI <7, than those with normal ALMI.

**Conclusion:** Visceral adiposity and a low lean muscle mass appear to be prevalent in South Asians with gender specific metabolic implications. Given the marked rise in CVD, diabetes and obesity related diseases in South Asia, urgent measures should be adopted to reduce the burden of illness due to these illnesses and diseases associated with frailty.

## Background

Cardiovascular diseases (CVD) resulting in stroke and ischaemic heart disease are the leading causes of death in Sri Lanka [1] and in South Asia [2]. The prevalence of CVD is predicted to increase by 109.o% from 2025 to 2050, with the crude mortality rates due to CVD predicted to increase by 85.3% in South Asia [3]. South Asia is expected to have the highest age-standardised mortality due to CVD (141 deaths per 100,000 population), by 2050 [3]. Obesity is an important risk factor for CVD and an individual in classified as overweight or obese based on an individual body mass index (BMI) [4]. Due to the differences in the body composition, the BMI cut-off values that define overweight and obesity are different for South Asian populations compared to individuals of Caucasian ethnicity [4]. However, the BMI has many limitations, as it is affected by the muscle mass of an individual and does not indicate the distribution of adiposity.

There is a marked increase in obesity and obesity related disorders in South Asia and Southeast Asia [5]. Obesity is associated with an increase in for developing CVDs, diabetes, cancers, obstructive sleep apnoea, dementia osteoarthritis and also is an important risk factor for development of severe disease in many infections [6-8]. However, there are many phenotypes of obesity and the distribution of fat, i.e. truncal vs subcutaneous fat, is associated with higher risk of CVD and many other diseases [9]. Furthermore, visceral adiposity when associated with a lower muscle mass (sarcopenic obesity), is associated with a higher risk of CVD and other metabolic diseases compared to those with a similar BMI, but a high muscle mass [9, 10]. Overweight individuals who have a higher amount of subcutaneous fat compared to visceral fat, are shown to have a lower risk of developing CVD [11]. Obese or overweight individuals, as defined by higher BMI, who were younger with lower visceral fat and physically active are ‘metabolically healthy’ and are at lower risk of developing CVD [12, 13]. Adipose tissue distribution has shown to vary among different ethnic groups, with the varying risks of developing CVD [12]. Although the waist circumference has been shown to correlate well with visceral adiposity, it cannot differentiate between visceral adiposity and subcutaneous adiposity [11, 12]. In addition to distribution of adipose tissue reduced skeletal muscle mass and strength has shown to be an important risk factor for CVD and diabetes [14]. Individuals with high visceral fat and lower lean muscle mass and strength are considered to have sarcopenic obesity [14]. Redistribution of fat during aging along with sarcopenia leads to ‘inflammaging’ which is a chronic low grade inflammatory state, which results in an increased risk of developing CVD, type 2 diabetes, cancers and other obesity related complications [15].

There is wide variation in sarcopenia and fat distribution among different ethnic groups [16]. Further, due to these differences in lean muscle mass and adipose tissue mass and distribution among different ethnic groups, the classifications used to define excess visceral adipose tissue, lean muscle mass and sarcopenia may need to be adjusted to different ethic groups, to evaluate risks for developing CVD, diabetes and other obesity related diseases. For instance, different BMI cut-off values and waist circumference cut-off values are used for South Asians for defining overweight, obesity and the presence of central obesity [17]. Currently, there is limited data of distribution of adiposity, lean muscle mass and metabolic risk factors and inflammatory mediators in South Asian populations, living in those countries and following dietary and lifestyle patterns in those countries. As the South Asian population currently constitutes 25% of the world population and given the marked rise of diabetes, CVD, obesity among South Asians, it would be important to identify the adiposity and lean muscle mass in healthy South Asians and those with metabolic diseases, to understand distribution of these parameters in these populations.

The measurement of adiposity and lean muscle indices by dual-energy X-ray absorptiometry (DEXA) is considered to be comparable to MRI and is currently the most widely used, and reliable method to assessing the body composition [18]. Therefore, we sought to assess the adiposity and lean muscle mass indices, along with metabolic risk factors and inflammatory mediators in a South Asian population to understand how these parameters associate with metabolic disease.

## Methods

### Study participants and screening for the presence of metabolic disease

We recruited 139 individuals by open advertisement in Sri Lanka, aged ≥20 years following informed written consent. Anthropometric measurements were taken, and fasting blood samples obtained to determine their metabolic profile. The lipid profile and fasting blood sugar (FBS) were analysed using the fully automated biochemistry analyser (Thermo scientific Indiko) in blood samples obtained following 10 hours of fasting. Levels of Apolipoproteins A1 and B were measured in serum stored at -80 C, using ApoA1 and ApoB quantitative ELISA (Mabtech, Germany) following manufacturer’s guidelines provided.

The presence of metabolic disease was defined based on criteria laid down by the International Diabetes Federation Task Force on Epidemiology and Prevention [19]. Accordingly, those with a waist circumference over the population cut-off values (≥80cm for females and ≥90cm for males), triglycerides ≥150 mg/dL (1.7 mmol/L), an HDL <40 mg/dL (1.0 mmol/L) in males; <50 mg/dL (1.3 mmol/L) in females, a systolic blood pressure of ≥130 and/or diastolic blood pressure of ≥85 mm Hg, or a FBS of ≥100 mg/dL [19].

### Ethical considerations

Ethical approval was obtained from the Ethics Review Committee of the Faculty of Medical Sciences, University of Sri Jayewardenepura, Sri Lanka (62/19). Informed written consent was obtained from all the recruited individuals. Samples were collected and stored according to the ethical regulations.

### Anthropometric measures

Individuals were weighed on a digital scale with an accuracy of ±0.1kg with the capacity to measure up to 150kg, while wearing light clothing and barefooted. The height was measured barefooted, using a stadiometer with an accuracy of ±0.1cm, with the heel touching the wall, feet flat and the head straight. Waist circumference was measured using a non-stretchable tape between the lowest rib and the iliac crest after an exhalation. Hip circumference was measured around the widest lateral extension of the pelvis. As the population was of South Asian ethnicity, those with a BMI below 18.5 kg/m^2^ were classified as underweight, 18.5-23.8 kg/m^2^ normal weight, 23.9-27.5 kg/m^2^as overweight and above 27.5 kg/m^2^ as obese [17].

### Dual energy Xray absorptiometry (DEXA) scanning to determine body composition

Hologic Discovery W DEXA scanner configured with software version 12.1 was used to conduct full body DEXA to assess bone mineral density, total body fat percentage, fat mass index, total lean mass and appendicular lean mass, visceral adipose tissue (VAT) area. The scans were performed by a trained individual and calibrations were conducted following the National Health and Nutrition Examination Survey, Body Composition Analysis (NHANES BCA) protocol, to ensure the measurements are consistent and accurate [20]. The total body fat percentage was calculated as the percentage of fat mass of the total body fat mass, while the visceral adipose tissue (VAT) area was measured by drawing a line in the muscle wall surrounding the abdominal cavity. Other definitions and how the calculations of the different parameters are indicated below.

### Fat mass index (FMI)

Fat mass index assesses the body fat mass relative to height providing a more specific indication of the degree of adiposity in the body. [20]. associated with increased risk of developing metabolic diseases and cardiovascular diseases. FMI was calculated using the formula.

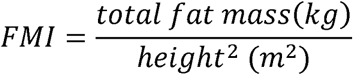

Based on the FMI, individuals were classified as having a low, normal or high FMI. Accordingly, females, with a FMI of <5 kg/m^2^ were considered as a low FMI, 5-9 kg/m^2^ normal and > 9 kg/m^2^ as high. Males with a FMI of <3 kg/m^2^ were considered as having a low FMI, 3-6 kg/m^2^ normal and > 6 kg/m^2^ as high FMI [21].

### Appendicular lean mass index (ALMI)

ALMI is measured to evaluate the skeletal muscle mass in the limbs relative to height [22]. The measurement is particularly important in identifying sarcopenia along with assessments of strength [23]. ALMI was calculated using the formula and An ALMI of <5.5 was considered to indicate sarcopenia in females and a values <7 indicative of sarcopenia in males [24].

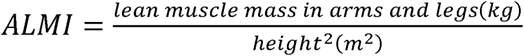

### Skeletal muscle index (SMI)

Skeletal muscle mass index (SMI) was also evaluated in individuals, as it is a more reliable indicator of the lean muscle mass, as the ALMI under-estimates the presence of low muscle mass in obese and overweight individuals.

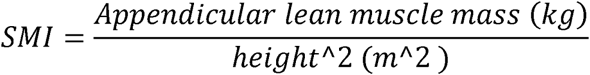

### Fat mass ratio (trunk to leg fat mass ratio)

The trunk-to-leg fat mass ratio assesses fat distribution, with a higher ratio indicating central adiposity linked to metabolic and cardiovascular risks.

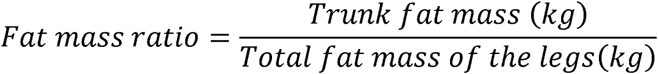

### Statistical analysis

Statistical analysis was performed using GraphPad Prism version 10.2.3. As the data were not normally distributed, (as determined by the frequency distribution analysis of GraphPad Prism) nonparametric statistical methods were used for the analysis of data. Mann Whitney U test (two tailed) was used to compare median values of metabolic markers with interquartile ranges between normal vs high VAT area, FMR and FMI groups, sarcopenic vs non-sarcopenic groups, and those with vs without predicted sarcopenic obesity. Spearmon correlation coefficients were calculated to assess the strength and direction of linear relationships between variables, such as waist circumference and VAT mass and VAT area correlations. The receiver operator curves (ROCs), were used to calculate the area under the curve (AUC), values and sensitivity and specificity of different cut-off values of VAT area and VAT mass that associate with metabolic risk factors, using the Wilson/Brown method.

## Results

### Anthropometric measurements in the study population

52 (37%) of the individuals were females and 87 (63%) males. 32 (62%) of the females and 65 (75%) of the males had at least one metabolic risk factor defined by the International Diabetes Federation Task Force on Epidemiology and Prevention [19]. The basic demographic details and metabolic risk factors of the study population is shown in table 1.

**Table 1:** Demographic and laboratory characteristics with anthropometric measures of the study population.

| Characteristic | Females<br>N (%) | Males<br>N (%) |
| --- | --- | --- |
| Age (mean, ± SD) | 38.5±10.8 | 39.7±10.7 |
| FBS >100mg/dL | 5 (10%) | 23 (26%) |
| Total cholesterol ≥200mg/dL | 29 (56%) | 50 (57%) |
| HDL (≤40mg/dL in males and ≤50mg/dL in females) | 21 (40%) | 31 (36%) |
| LDL ≥100 mg/ dL | 42 (81%) | 76 (87%) |
| Triglycerides $\geq 150\text{mg/dL}$ | 0 | 16 (18%) |
| Waist circumference<br>($\geq 80\text{cm}$ in females and $\geq 90\text{cm}$ in males) | 19 (37%) | 44 (50%) |
| BMI |  |  |
| $< 18.5 \text{ kg/m}^2$ | 2 (4%) | 3 (3%) |
| $\geq 23.9 \text{ kg/m}^2$ | 20 (38%) | 49 (56%) |

### Relationship between visceral fat and metabolic risk factors

In previous studies, those with a VAT area of ≥100cm^2^ were at a higher risk of developing obesity related metabolic disorders [25, 26]. Therefore, we evaluated the cardiometabolic risk factors and inflammatory mediators in those with a high VAT area (≥100cm^2^), compared to others. 23(44%) females and 67(77%) males had a VAT area of ≥ the cut-off value. Females with a VAT area ≥ 100 cm^2^, were significantly likely to be older, have significantly lower HDL and ApoA1 levels and significantly higher FBS levels (table 2). In males who had a VAT area of ≥ 100cm^2^, there was no difference in age, or any metabolic parameters or inflammatory mediators.

**Table 2:** Association of metabolic risk factors VAT area in males and females.

|  | Females |  | P value | Males |  | P value |
| --- | --- | --- | --- | --- | --- | --- |
| | VAT $< 100$ | VAT $\geq 100$ | | VAT $< 100$ | VAT $\geq 100$ | |
| Age | 36±10 | 41.4±10.9 | 0.028 | 37.1±11.4 | 40.5±10.3 | 0.16 |
| HDL | 59.5 (47.25- | 49.5 (40- | 0.03 | 44 (37-56) | 42 (36-53) | 0.9 |
|  | 77.25) | 60.75) |  |  |  |  |
| LDL | 133.5 (107.5-146.3) | 122.2 (100.5-159.8) | 0.5 | 133 (119-167) | 136 (117-166.3) | 0.9 |
| Total Cholesterol | 208.5 (186-231.8) | 192 (169.8-213.8) | 0.1 | 195 (175-235) | 209.5 (197.5-236) | 0.3 |
| FBS | 85.5 (84-91) | 90 (86.25-98.5) | 0.008 | 89 (86-97) | 91 (86-102.8) | 0.2 |
| ApoA1 | 163.5 (128.3-198.5) | 140 (117-163) | 0.04 | 130 (105-159) | 126 (108.5-150) | 0.9 |
| ApoB | 78.5 (72.75-101) | 80 (73-102) | 0.9 | 102 (75-120) | 95 (84.5-113) | 0.8 |
| ApoB/Apo A1 ratio | 0.53 (0.38-0.82) | 0.62 (0.43-0.87) | 0.2 | 0.85 (0.64-1.14) | 0.81 (0.59-0.97) | 0.8 |
| IL-6 | 12.48 (6.52-23.25) | 16.7 (8-41) | 0.1 | 15.2 (12.48-27) | 12.48 (6.1-23.35) | 0.1 |
| IL-1β | 4.2 (2.85-5.95) | 4.2(2.7-5.3) | 0.7 | 4.19 (2.7-5) | 3.9 (3-12.6) | 0.8 |

### Cut-off values for visceral fat area and waist circumference associated with increase in the prevalence of the obesity-related disorders in South Asian populations

Although previous studies have suggested a VAT are cut-off value of ≥100cm^2^ as a risk factor of obesity related diseases in previous studies, in Southeast Asian populations (Koreans), a values of >103.8 cm2 was found to have a sensitivity 74.5% and a specificity 64.7% being associated with an increased risk for metabolic diseases [25, 26]. Therefore, we proceeded to determine the VAT area cut-off values in our population that associated with the presence of metabolic risk factors. The differences in the VAT area was compared in those who had at least one of the metabolic risk factors defined by the International Diabetes Federation Task Force on Epidemiology and Prevention [19], and the receiver operator curves (ROCs), were calculated to define potential cut-off values for the VAT area that associated with the presence of metabolic risk factors. The VAT areas although higher in males with metabolic risk factors, did not achieve significance (p=0.056),), with the area under the curve (AUC) value being 0.63. A VAT area ≥ 99.8 cm^2^ was associated with an 80% sensitivity and 27.2% of specificity of presence of a metabolic risk factor. However, in females the VAT areas were significantly higher in those with a metabolic risk factor (p=0.04), with an AUC value of 0.67.

As the VAT area did not appear to correlate well with the metabolic risk factors, we also analysed the association of the VAT mass with metabolic risk factors. Although the VAT mass was high in males with metabolic risk factors, this was not significant (p=0.056), with the area under the curve (AUC) value being 0.63. In females, similar values were obtained with the VAT mass being significantly higher in those with a metabolic risk factor (p=0.04), with an AUC value of 0.67.

As VAT area appeared to associate with FBS, and since VAT area did not associate with all metabolic risk factors, we proceeded to define cut-off values that associated with a FBS ≥100mg/dl in both males and females. The VAT area was significantly higher in males (p=0.0005, AUC=0.75) and females (p=0.0005, AUC=0.97) with a FBS of ≥100mg/dl. In males a VAT area of ≥145 cm^2^ had a sensitivity of 78.3% and a specificity of 75% with a FBS of ≥100mg/dl and a likelihood ratio of 3.1 (Fig 1A). In females a VAT area of ≥130.5 cm^2^ had a sensitivity of 100% and a specificity of 93.62% with a FBS of ≥100mg/dl (Fig 1B) and a likelihood ratio of 15.6. We also compared the waist circumference with the VAT area, which showed a significant correlation in males (Spearman’s r=0.78, p<0.0001) and females (Spearman’s r=0.77, p<0.0001). In males a waist circumference of <90.5cm had a sensitivity of 62.5% and a specificity of 73.9% with a FBS of ≥100mg/dl, with a likelihood ratio of 2.39. In males a waist circumference of <90.5 had a sensitivity of 100% and a specificity of 68.1% with a FBS of ≥100mg/dl, with a likelihood ratio of 3.1.

**Figure 1.**
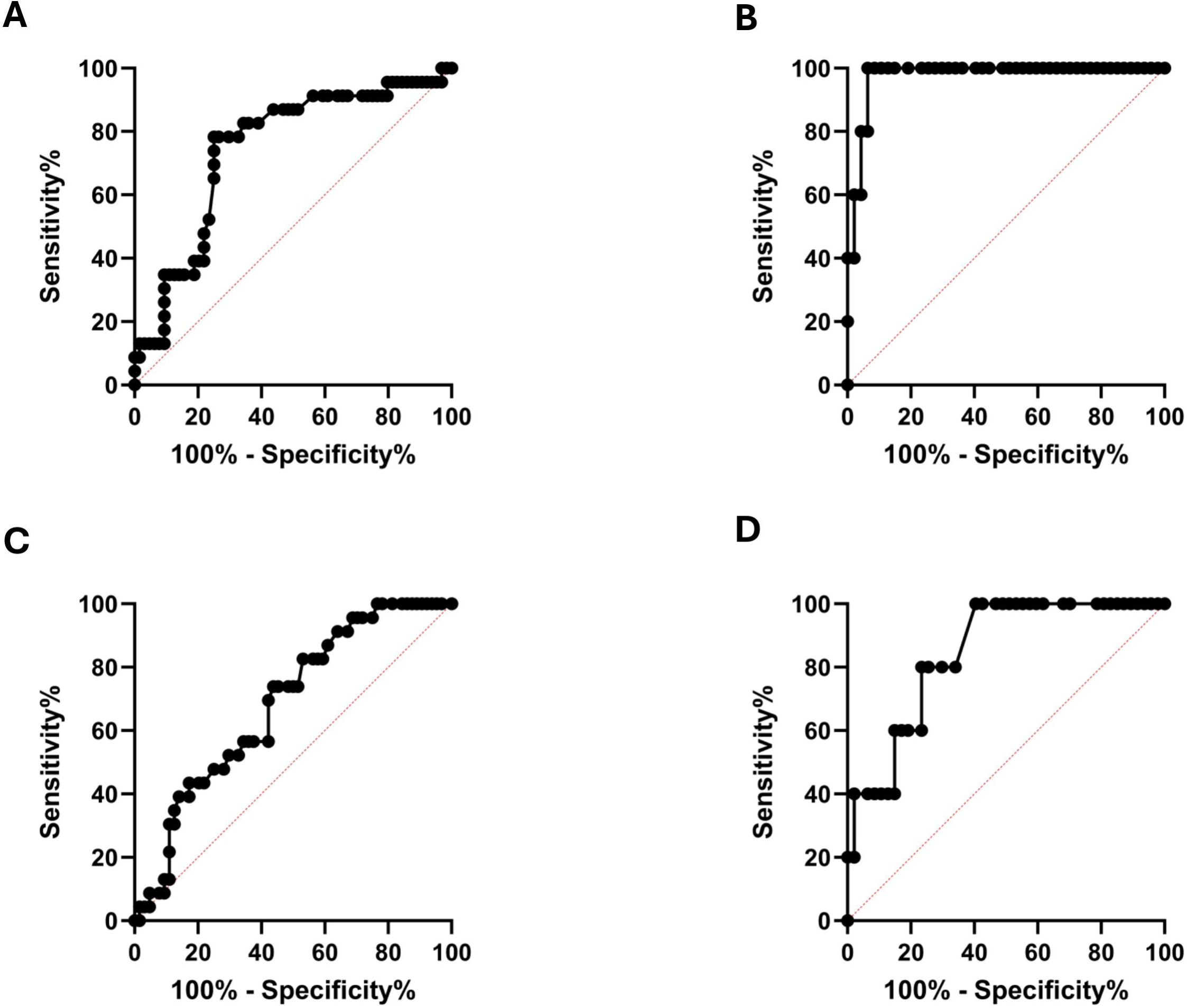
ROC curves for VAT area and FMR in predicting elevated FBS in Sri Lankan adults. Receiver operating characteristic (ROC) curves illustrating the diagnostic performance of visceral adipose tissue (VAT) area and fat mass ratio (FMR) in predicting elevated FBS in Sri Lankan males (n=87) and females (n=52). (A) VAT area in males: AUC=0.5, p=0.0005; cut off ≥145cm^2^ yielding 78.3% sensitivity, 5% specificity and a likelihood ratio of 3.1. (B) VAT area in females: AUC=0.9, p=0.0005; cut off ≥130.5cm^2^ with 100% sensitivity, 93.6 specificity, and a likelihood ratio of 15.6. (C) FMR in males: AUC=0.68, p=0.009; cut off ≥1.61 achieving 73.9% sensitivity, 56.2% specificity, and a likelihood ratio 1.69. (D) FMR in females: AUC=0.84, p=0.01; cut off ≥1.32 achieving 80% sensitivity, 76.6% specificity, and a likelihood ratio of 3.4.

### Association of metabolic risk factors with other adiposity indices

The fat mass index (FMI) as it assesses the body fat mass relative to height providing a more specific indication of the degree of adiposity in the body. To understand the relationship with FMI and metabolic risk factors, we compared the metabolic and inflammatory markers in females and males with an FMI above and below the cut-off value. 34 (65%) females had an FMI above the cut-off value (≥9), while 79 (91%) males had an FMI above the cut-off value (≥6). There were no significant differences in the age in females and males with an FMI above the cut-off value. Females with an FMI of ≥9 had significantly higher FBS levels (table 3). There were no differences in metabolic risk factors or inflammatory mediators in males with an FMI above the cut-off value (≥6).

**Table 3:** Association of metabolic risk factors with FMI and fat mass ratio in males and females of South Asian origin.

| **Association of metabolic risk factors with Fat mass index (FMI)** |  |  |  |  |  |  |
| --- | --- | --- | --- | --- | --- | --- |
|  | Females |  | P value | Males |  | P value |
|  | FMI<9 | FMI ≥9 |  | FMI <6 | FMI ≥6 |  |
| Age | 36.2±8.8 | 39.7±11.3 | 0.1 | 41.4±11.7 | 39.6±10.6 | 0.5 |
| HDL | 59.5(47.5-82) | 53 (41-64) | 0.3 | 40.5 (35-53) | 45 (37-54) | 0.2 |
| LDL | 131.5(93.25-150.5) | 129 (104.3-159.3) | 0.9 | 121.5 (112-160.5) | 136 (118-167) | 0.3 |
| Total | 204.5 | 199.5 (175- | 0.5 | 182 (160.5- | 208 (189- | 0.09 |
| Cholesterol | (173-231.5) | 221.8) |  | 225.3) | 237) |  |
| FBS | 85 (81.75-90.5) | 89.5 (85-95.25) | 0.01 | 87 (80.75-91.25) | 91 (87-102) | 0.09 |
| ApoA1 | 158 (135.8-201.3) | 144 (120-172.5) | 0.2 | 115.5 (105-154.5) | 126 (109-150) | 0.5 |
| Apo B | 78(72.25-103.5) | 80(73-99.5) | 0.8 | 86 (71.25-138.5) | 96.5 (84.25-115.8) | 0.5 |
| ApoB/ApoA<br>1 ratio | 0.53(0.33-0.79) | 0.62 (0.42-0.82) | 0.4 | 0.81 (0.48-1.15) | 0.82 (0.6-0.98) | 0.8 |
| IL-6 | 14.5 (8-25.5) | 14.5 (7-26.25) | 0.9 | 14.5 (8.93-20.68) | 13.44 (7-24) | 0.9 |
| IL-1β | 4.2 (3-8.1) | 4.2 (2.7-5.5) | 0.3 | 4.3 (3-5.2) | 3.9 (3-5.6) | 0.8 |
| BMI | 21(20-23) | 24(22-26) | <0.0001 | 19.5(18.25-21) | 25(23-28) | <0.0001 |
| Waist<br>circumference | 70.5(65.5-74) | 81(74-84) | <0.0001 | 78(74-87.5) | 91(85-96) | 0.0014 |
| **Association of metabolic risk factors with fat mass ratio** |  |  |  |  |  |  |
|  | Females |  | P value | Males |  | P value |

| Median,<br>(IQR) | FMR <1.2 | FMR ≥1.2 |  | FMR <1.7 | FMR ≥ 1.7 |  |
| --- | --- | --- | --- | --- | --- | --- |
| Age | 35.7±9.5 | 40.6±11.1 | 0.05 | 38.9±12.2 | 41.1±7.5 | 0.4 |
| HDL | 57 (488-74.25) | 52 (41-62.25) | 0.3 | 43 (36-56) | 45 (37.5-53.5) | 0.5 |
| LDL | 121.5 (98-143.3) | 133 (110.8-160.3) | 0.3 | 136 (117.8-174) | 136 (118-160) | 0.7 |
| Total<br>Cholesterol | 201 (173.5-223.3) | 203.5 (175-231.3) | 0.6 | 204 (183-241.3) | 208 (195.5-231) | 0.8 |
| FBS | 85 (83.5-91) | 89 (85.75-96.25) | 0.01 | 90.5 (86.75-99) | 91 (86-110) | 0.4 |
| ApoA1 | 162.5 (133.8-199.8) | 140 (120-170) | 0.1 | 127.5 (103-153.8) | 124.5 (113-146.3) | 0.7 |
| ApoB | 76 (67.75-87.5) | 92 (75-103) | 0.03 | 101 (84.5-118.5) | 90.5 (77.25-107) | 0.2 |
| ApoB/ApoA<br>1 ratio | 0.47 (0.34-0.78) | 0.63 (0.47-0.87) | 0.03 | 0.86 (0.63-1.1) | 0.75 (0.6-0.89) | 0.1 |
| IL-6 | 22.5 (6.5-45) | 13.8 (7.54-<br>) | 0.2 | 13.1 (6.28-22.5) | 15.16 (8.85-32.03) | 0.1 |
|  |  | 19.5) |  |  |  |  |
| IL-1β | 4.5 (3-5.95) | 3.6 (2.7-5.3) | 0.1 | 4.1(3-5.3) | 3.76 (2.63-5.658) | 0.4 |
| BMI | 23.5(20.75-26) | 23(22-25) | 0.6 | 23(22-27) | 25(23.5-28) | 0.06 |
| Waist circumference | 73.5(66-79) | 81(74-83.25) | 0.003 | 88(80-95) | 93(87-98.5) | 0.002 |

The fat mass ratio (FMR) has been used to diagnose lipodystrophy in HIV patients on anti-retroviral therapy and the cut off values indicating lipodystrophy in different populations vary from 1.5 in Caucasian males in France [27] and 1.96 in Portuguese men and 1.33 in Portuguese women [28]. Recently, the FMR has also shown to associate with metabolic risk factors and biomarkers and was found to associate with diabetes in the absence of obesity [13]. Therefore, based on the recent large study carried out in the UK Biobank study participants, we used a FMR ≥1.7 in male participants and ≥1.2 in female participants to indicate potential lipodystrophy [13]. 30 (58%) females had an FMR above the cut-off value (≥1.2), while 33 (38%) males had an FMR above the cut-off value (≥1.7).

The metabolic characteristics and inflammatory mediators of those with a high FMR is shown in table 3. There was no difference in the age in females or males with an FMR above the cut-off value that indicated lipodystrophy. Females with an FMR of ≥ of 1.2 were had significantly higher FBS and ApoB levels with significantly lower ApoA1 levels (table 3). There were no differences in metabolic risk factors or inflammatory mediators in men with a FMR of ≥ 1.7.

As the cut-off values defined by Agrawal et al did not appear to associate with any metabolic derangement in males, we sought to identify FMR cut-off values for males and females in the Sri Lankan population, that may associate a FBS of ≥100mg/dl. The FMR was significantly higher in males (p=0.009, AUC=0.68) and females (p=0.01, AUC=0.84) with a FBS of ≥100mg/dl. In males a FMR of ≥ 1.61 had a sensitivity of 73.9% and a specificity of 56.2% with a FBS of ≥100mg/dl, with a likelihood ratio of 1.69 (Fig 1C). In females a FMR of ≥1.32 had a sensitivity of 80% and a specificity of 76.6% with a FBS of ≥100mg/dl, with a likelihood ratio of 3.4 (Fig 1D).

### Skeletal muscle mass and related indices in females and males of South Asian origin

ALMI is measured to evaluate the skeletal muscle mass in relation to the height and is one of the parameters considered in the diagnosis of sarcopenia along with grip strength. 34 (65%) females had an ALMI below the cut-off value (<5.5) associated of sarcopenia, while 47 (54%) males had an ALMI below the cut-off value (<7). The metabolic and inflammatory markers were not different females with an ALMI of above the cut-off value compared to those with lower values. However, in males the ApoB levels, ApoB/ApoA1 ratio and serum cholesterol were significantly higher in males with a lower ALMI than those with an ALMI above the cut-off value (table 4).

**Table 4:** Association of metabolic risk factors with ALMI in males and females of South Asian origin.

|  | Females |  | P value | Males |  | P value |
| --- | --- | --- | --- | --- | --- | --- |
| Median, (IQR) | ALMI<5.5 | ALMI≥5.5 | | ALMI<7 | ALMI≥7 | |
| Age | 38.6±11.7 | 38.4±8.7 | 0.9 | 39.7±12.2 | 39.8±8.6 | 0.9 |
| HDL | 60 (44-75.75) | 50 (40.5-58.25) | 0.053 | 45 (38-56) | 43 (36-52.75) | 0.4 |
| LDL | 126 (99.5-159.5) | 130.5 (105.8-148) | 0.5 | 136 (115-153) | 136 (118.3-180.8) | 0.2 |
| Total Cholesterol | 204.5 (175.8-230.3) | 194.5 (171.3-218.3) | 0.3 | 199 (180-222) | 215.5 (193.5-245.3) | 0.04 |
| FBS | 89.5 (85- | 87 (84-91) | 0.2 | 91 (86-99) | 90 (86- | 0.9 |
|  | 94) |  |  |  | 106.3) |  |
| ApoAI | 161<br>(122.3-189) | 140 (120-161.5) | 0.1 | 126 (111-160) | 122 (104-144) | 0.2 |
| ApoB | 77.5<br>(72.25-101) | 80 (73-101.5) | 0.8 | 90(74-112) | 103 (88.5-121) | 0.04 |
| ApoB/ApoAI<br>ratio | 0.56<br>(0.39-0.83) | 0.62 (0.52-0.83) | 0.3 | 0.76 (0.49-0.95) | 0.87 (0.68-1.13) | 0.03 |
| IL-6 | 13.14<br>(6.78-24) | 15.6 (9.31-45.88) | 0.3 | 13.8 (8.89-23.55) | 12.48 (6-31.6) | 0.7 |
| IL-1β | 4.35 (3-5.3) | 3.61 (2.7-5.75) | 0.7 | 3.9(3-5.3) | 4.19 (2.39-5.72) | 0.8 |
| BMI | 22(21-24) | 25(23-27) | 0.0031 | 23(22-24) | 26.5(25-29) | <0.0001 |
| Waist<br>circumference | 74(70-81) | 78.5(73.75-84.5) | 0.12 | 86(80-92) | 93(88.25-101) | 0.0001 |

### Sarcopenic obesity and metabolic and inflammatory risk factors

As we did not measure that handgrip strength of any of the study participants, we considered females who had a ALMI below the cutoff value (≤5.5) and FMR above the cutoff value (≥1.2) potential of sarcopenic obesity (SO), while in males had ALMI below the cutoff value (≤7) and FMR above the cutoff value (≥1.7) were considered to have potential SO. 22 (42%) females potential SO, while 13 (15%) had potential SO. There were no differences in metabolic risk factors or differences in levels of inflammatory mediators (table 5).

**Table 5:** Metabolic risk factors and inflammatory markers in those with and without sarcopenic obesity.

|  | Females |  | P value | Males |  | P value |
| --- | --- | --- | --- | --- | --- | --- |
| Median, | Potential | Potential |  | Potential | Potential |  |

| (IQR) | Sarcopenic obesity+ | Sarcopenic obesity - |  | Sarcopenic obesity+ | Sarcopenic obesity - |  |
| --- | --- | --- | --- | --- | --- | --- |
| Age | 40.5±12.2 | 37±9.3 | 0.2 | 41.8±7 | 39.4±11.2 | 0.3 |
| HDL | 56.5<br>(40.75-68) | 53.5 (46-68.5) | 0.9 | 45 (41-51.5) | 42 (36-53) | 0.6 |
| LDL | 131.5<br>(105.3-161) | 128 (101.5-143.3) | 0.7 | 136 (119-148.5) | 136 (117.8-173.3) | 0.4 |
| Total Cholesterol | 204 (175-231.3) | 197.5 (174.3-223.3) | 0.6 | 201 (189.5-220) | 211 (182-242) | 0.4 |
| FBS | 88.5 (85-114) | 87 (84-91.25) | 0.1 | 87 (84.5-105) | 91 (87.75-101.3) | 0.5 |
| ApoA1 | 142.5(114-173.8) | 153 (129.5-183.5) | 0.5 | 126 (120-155) | 126 (105-150) | 0.5 |
| ApoB | 91 (75-123) | 77 (68.5-99.5) | 0.1 | 95 (72.5-102) | 96 (84-118) | 0.3 |
| ApoB/ApoA1 ratio | 0.63<br>(0.46-0.9) | 0.54 (0.4-0.78) | 0.1 | 0.73 (0.56-0.83) | 0.84 (0.6-1) | 0.1 |
| IL-6 | 12.4 (7.5-18.8) | 15.4 (6.1-29) | 0.2 | 13.8 (6.9-24) | 13.3 (7.2-22.5) | 0.7 |
| IL-1β | 3.9 (2.7- | 4.2 (3-5.6) | 0.4 | 3.6(3-5.1) | 5.7 (4.9- | 0.7 |
|  | 5.3) |  |  |  | 12.6) |  |
| BMI | 22(21-24) | 25(23-27) | 0.0031 | 24(22-24.5) | 24.5(22-28) | 0.15 |
| Waist<br>circumference | 74(70-81) | 78.5(73.75-84.5) | 0.1 | 91(86.5-95.5) | 89.5(83.75-96) | 0.7 |

Potential sarcopenia obesity (SO) was defined as ALMI below the cutoff value (≤5.5) and FMR above the cutoff value (≥1.2) for females and an ALMI below the cutoff value (≤7) and FMR above the cutoff value (≥1.7) for males.

Serum IL-6 level inversely correlated with HDL (Spearmans’r=-0.26, p=0.003), ApoAI (Spearmans’r=-0.21, p=0.02), but also with ApoB (Spearmans’r=-0.27, p=0.001). Serum IL-1β levels did not correlate with any of the metabolic or anthropometric parameters. Interestingly, ApoAI inversely correlated with the ALMI values (Spearmans’r=-0.33, p<0.0001) and with the waist circumference (Spearmans’r=-0.29, p=0.0008) and the VAT area (Spearmans’r=-0.19, p=0.03). ApoB was found to correlate with the ALMI (Spearmans’r=0.30, p=0.0005) and FMR (Spearmans’r=0.30, p=0.002).

## Discussion

In this study, we have assessed the adiposity and lean muscle indices of Sri Lankan females and males and have compared them with conventional anthropometric measures such as the BMI and waist circumference. Further, we proceeded to investigate the association between VAT, distribution of adipose tissue preferentially in truncal areas instead of limbs (FMR) and lean muscle mass induces with metabolic risk factors and inflammatory mediators. We found that females with high VAT area and a FMR had significantly higher FBS and lower ApoA1 and HDL levels. However, there were no differences in metabolic risk factors or inflammatory mediators in males who had a VAT area or an FMR above the cut-off value. Instead, men with lower skeletal mass (an ALMI < the cut-off values) had significantly higher cholesterol and ApoB levels, with an increase in ApoB/ApoA1 ratios, which are important risk factors for CVD [29]. In contrast, women who had an ALMI below the cut-off values did not have an increase in any of the metabolic risk factors or inflammatory mediators. The reasons for adiposity indices in women significantly associating metabolic risk factors, but not in men, while the lean muscle indices associating with dyslipidaemia in men but not in women is not clear. Based on the data, it appears that the lean muscle mass is likely to play a more significant role in reducing the risk of metabolic disease in males than in females, which should be further investigated.

Although healthy individuals in the community were recruited from open advertisement, 62% of the females and 75% of the males had at least one metabolic risk factor defined by the International Diabetes Federation Task Force on Epidemiology and Prevention [19]. Further, 44% of females and 77% of males had a VAT area of ≥ 100cm^2^, 65% females and 91% males had an FMI of above the cut-off value and 58% females, and 38% males had an FMR above the cut-off value. Therefore, this data shows the prevalence of obesity and prevalence of individuals with maldistribution of fat in South Asian populations. Given that the mean age of females was 38.5 years, while the mean age of males was 39.7 years, this highlights the population at risk of developing CVD, diabetes, cancers and other obesity associated diseases in future. Given that CVD diseases is currently the leading cause of death in Sri Lanka [1] and in South Asia [2], extensive public health programs would be required to reduce the burden of these diseases in these countries. Furthermore, low grade inflammation and insulin resistance is also associated with an increased risk of breast, colorectal, endometrial, pancreatic, prostate cancer and many haematological malignancies [30]. Therefore, given the marked increase in these diseases associated with an increase in visceral adiposity, urgent measures are required to advocate healthy lifestyle, exercise and healthy eating habit for prevention of these diseases.

65% females and 54% males had an ALMI below the cut-off value. Although a lower ALMI was not associated with an increase in metabolic risk factors in females, a lower ALMI is associated with reduction in strength, aerobic capacity ultimately resulting in a reduction in functional capacity [31]. Although we did not test the muscle strength in the study population, a reduced lean muscle mass in younger adults or middle-aged adults is likely to lead to an increase in frailty as individuals age. The reasons for most individuals in this study having a lean muscle mass, below the range associated with sarcopenia could be due to multiple factors such as lack of any physical activity or due to ongoing chronic inflammation and insulin resistance [31]. The reduced lean muscle mass in individuals will further lead to a reduction in functional capacity, thereby further limiting the ability to engage in physical activity. This in turn, is likely to lead to a viscous cycle of further increase in visceral adiposity, reduced muscle mass and increased risk of all obesity related diseases. In addition, although we did not see any differences in metabolic risk factors or inflammatory mediators in females and males with potential SO, it would be important to carryout large longitudinal studies to identify cut-off values for FMR and ALMI values that would be more suitable for South Asian populations, in order to carry out required interventions.

## Conclusions

In summary, we found that 44% of females and 77% of males had high amount of visceral adiposity, with 91% males with a fat mass above cut-off values. Further, while 58% females, and 38% males, had maldistribution of adiposity with increase in fat in their trunk vs limbs, 65% females and 54% males also had lean muscle mass, in the sarcopenic range. Given the marked rise in CVD, diabetes and obesity related diseases in South Asia, urgent measures should be adopted to reduce the burden of illness due to these illnesses and diseases associated with frailty.

## Abbreviations

ALMI: Appendicular lean mass index
AUC: Area under the curve
BMI: Body mass index
CVD: Cardiovascular diseases
DEXA: Dual energy X-ray absorptiometry
FMI: Fat mass index
FMR: Fat mass ratio
NHANES BCA: National Health and Nutrition Examination Survey, Body Composition Analysis
ROC: Receiver operator curves
SO: Sarcopenic obesity
VAT: Visceral adipose tissue

## Declarations

### Ethics approval and consent to participate

Ethical approval was obtained from the Ethics Review Committee of the University of Sri Jayewardenepura. All participants gave informed written consent.

### Consent for publication

Not applicable.

### Availability of data and material

All data is available within the manuscript.

## Funding

We are grateful to the University of Sri Jayewardenepura Sri Lanka (grant number ASP/01/RE/MED/2019/50).

### Authors’ contributions

Conceptualization: GNM, DNG, CJ

Data curation: DNG, JJ, PHC, TN, UA, AP

Experiments and investigations: DNG, JJ, PHC, TN, LP, UA, RHW, AP

Data analysis: GNM, DNG

Project administration and supervision: GNM, CJ

Funding acquisition: GNM, CJ

Writing the manuscript: DNG, GNM

Reviewing the manuscript: All authors read and approved the final manuscript

## Data Availability

All data produced in the present work are contained in the manuscript.

